# A deeper look at COVID-19 CFR: health care impact and roots of discrepancy

**DOI:** 10.1101/2020.04.22.20071498

**Authors:** Amirhoshang Hoseinpour Dehkordi, Reza Nemati, Pouya Tavousi

**Author notes:** Equal contribution (Reza Nemati, Pouya Tavousi).

## Abstract

Intensive care capacity and proper testing play a paramount role in the COVID-19 Case Fatality Rate (CFR). Nevertheless, the real impact of such important measures has not been appreciated due to the lack of proper metrics. In this work, we have proposed a method for estimating a lower bound for the number of positive cases by using the reported data on the oldest age group and the regions’ population distributions. The proposed estimation method improved the expected similarity between the age-distribution of positive cases and regions’ population. Further, we have provided a quantitative measure for the impact of intensive care on the critical cases by comparing the CFR among those who did and did not receive intensive care. Our findings showed that the chance of living among non-ICU receivers is less than half of ICU receivers (∼24% vs ∼60%).

## Introduction

Realistic modeling of COVID-19 behavior is essential for policy makers to put in place the most effective non-pharmaceutical interventions (NPIs) against this world-wide pandemic. In this regard, a key factor to assess the impact of NPIs on this pandemic is case fatality rate (CFR). Dictionary of Epidemiology defines CFR as “the proportion of cases of a specified condition that are fatal within a specified time” [1].

Ever since the COVID-19 outbreak started in China, several approaches have been practiced to correctly evaluate CFR. One of the earliest attempts to estimate the CFR for COVID-19 was done by Baud et al. [2]. They took into account a 14-day incubation time for COVID-19 and estimated mortality rates by dividing the number of deaths on a given day by the number of patients with confirmed COVID-19 infection 14 days before. However, the CFR calculated by Baud et al. was an overestimation due to under-testing at the beginning of the pandemic and improper phase lag estimation. Spychalski et al. estimated CFR based on the total number of confirmed cases and showed that this approach is least affected by the reporting biases [3]. In another study, Dehkordi et al. discovered that the distribution of daily confirmed cases and the number of deaths in a specific region have a quasi-linear relationship if an optimum phase lag for number of deaths is calculated. In their work, optimum phase lag was calculated by minimizing the error for the regression line while changing the phase lag. Then, CFR for that region was corrected by dividing the number of deaths on a specific date by the total number of confirmed cases on a shifted date (based on the optimum phase lag) Dehkordi et al. [4].

Regardless of the practiced method, CFR evaluation faces challenges that are manifested in the discrepancies in reported CFR values for different regions/countries. These discrepancies are generally known to appear because of two reasons: (1) inaccurate estimations of CFR; (2) the differences in health care quality metrics. However, the real effect of each factor has not been effectively quantified. As for the estimation of CFR, it must be realized that not all of the infected cases are symptomatic. For example, Liu et al. showed that 23% of transmission in Shenzen was originated from pre-symptomatic infections. Further, testing all members of society is not feasible at the moment due to limitation in throughput of current assays and lack of antibody detection tests ready for the public use. CFR can also be underestimated because of the delay between diagnosis and reporting of the positive cases [3]. As for the impact of health care, one must note that because of the nature of the disease, the intensive care capacity and the time that this cap is reached significantly influence the CFR. In light of this, there is a great need for quantitative assessment of the aforementioned factors on CFR.

The performance of any quantitative analysis, focused on CFR evaluations, is also influenced by our understanding of the mechanisms, by which the virus spreads. Ongoing research by the community has shed light on the reality of such mechanism. For example, it is now evident that respiratory, contact and aerosol transmission are the main mechanisms by which the virus spreads [5, 6]. Accordingly, countries that were heavily impacted by COVID-19 were forced to put in place various mitigation policies. Also, it has been shown that initial viral load is important for a more effective transmission of the disease [5, 7]. However, there is little known about whether people with different age have different susceptibility to the infection [8]. Given a higher mortality rate for older cases, in one study, Li et al. showed that more than 50% of early Patients with COVID-19 in Wuhan were more than 60 years old. However, under-representation of younger people was suspected to be attributed to the fact that some of young infected people were asymptomatic [5, 6, 9, 10]. Therefore, in the absence of a true picture of the disease behavior, data scientists are instead urged to practice realistic assumptions when evaluating CFR.

Herein, we attempt to show how demographical comparison of COVID-19 confirmed cases and population of different regions/countries can shed light on estimated CFR discrepancies for different regions. Also, in order to improve the estimation of number of positive cases, the data on the oldest age group, namely >80-year-olds, are used to calculate an infected population ratio. Such infected population ratio will then be used to estimate the number of positive cases in other age groups. Furthermore, using corrected estimated CFR (based on our previous work [4]), we provide a new perspective on the importance of healthcare system capacity. Importantly, a quantitative measure for comparing the access vs no access to ICU beds for critical cases, in terms of fatality rate, will be provided. In accomplishing so, the following assumptions are made: (1) Infection mainly occurs through the three aforementioned known mechanisms of transmission; (2) Susceptibility to catching the virus is not age-dependent; (3) On average, >80-year-old cases have less number of social contacts than younger population [11]; and (4) All >80-year-old cases are symptomatic and correctly identified. Because of our last two assumptions, our analysis provides only lower bound estimates to the positive number of cases.

## Method

The estimated CFRs were calculated using the regression method described in [4]. Briefly, an optimum phase lag for the number of deaths is calculated for which the distribution of daily confirmed cases and number of deaths have linear relationship. Optimum phase lag is calculated by minimizing an error for the regression line namely, absolute minimum error (MAE), while changing the phase lag. Using this regression method, optimum phase lags and in turn CFRs were calculated for different COVID-19 hotspots [4].

The data for the number of confirmed cases or deaths were taken from [12]. Also, the age-stratified data for confirmed cases were obtained from [13–16]. In order to improve the estimation of total number of COVID-19 confirmed cases, the following assumptions were made: (1) Susceptibility to infection, due to contact with a positive case, is independent of age; and (2) In estimating the number of positive cases, the level of infection is not taken into consideration; (3) Data on the oldest age group more accurately reflects the demography of COVID-19 due to the fact that the fraction of symptomatic cases over all positive cases is highest in this group; (4) all of the positive cases in the >80 age group are correctly identified and confirmed. Using the data for >80-year-old cases, corrected lower-bound estimates of positive cases were made for different hot spots, namely Lombardy, Italy and New York, USA. The distributions of estimated positive cases and confirmed cases, in regions with high and low CFRs, were compared against each other and against the population distribution, for South Korea, Germany, Italy and Spain.

The number of ICU beds and occupancy of such intensive cares were obtained from [17, 18]. The population of each region or country and mean free capacities were obtained from United Nations’ website [19], and Organisation for Economic Co-operation and Development (OECD) library [20], respectively. The impact of ICU on CFR was quantified by comparing the fatality rate between ICU-admitted vs non-ICU-admitted patients among critical cases. For this analysis, the CFR was monitored from the beginning of the outbreak in a specific region before and after the intensive health capacity cap was reached.

The following resources were used:

- hospital bed per person (HBPP) data available in [Roemer [17]], [Wikipedia [21]] and [worldbank [18]]
- geographical regions’ population data, available in [Nations [19]]; and
- mean health care free capacity data available in [OECD [20]]

## Results and Discussion

This article provides a quantitative measure for the impact of health care capacity on CFR. For this purpose, our previous method for CFR estimation [4] was leveraged to compare the fatality rate for critical cases who did and did not receive intensive care. Further, this work addresses the inconsistencies observed in COVID-19 CFR reported in different geographical regions.

### Lower bound estimation of positive cases

Establishing high-level health care policies rely on an accurate estimation of infected cases. Respiratory, contact and aerosol transmission are the main known mechanisms by which the virus spreads. It has also been shown that the susceptibility to the infection depends mainly on viral load and exposure to an infected person [5, 7]. Therefore, the age-distribution of infected cases is expected to be similar in shape to that of region’s population. However, we observe that, in regions with relatively higher CFR, the age-distribution of infected is skewed towards the older ages (Fig.2b,2a). Further, the observed differences between CFR of regions with similar population distributions are beyond differences in health care quality and other such contributing factors.

We hypothesize that these inconsistencies are related to how these testings are conducted and that the infection in older age groups is more symptomatic and thus more detectable. Although the asymptomatic positive cases, more prevalent among younger age groups, may go unnoticed, they equally contribute to the spread of virus in the society.

To address the aforementioned inconsistencies, the data on the oldest age group (i.e. >80-year-old group) are considered. Assuming that due to the severity of symptoms, all of the positive cases in this age group are identifiable, the ratio of positive cases over the entire population for this age group was calculated. Next, the calculated ratio was applied to the populations of other age groups to estimate a lower number for the number of positive cases for them. The reason that the obtained estimation is a lower bound of positive cases is two-fold:

1. The method assumes that all of the infected cases in the >80 age group are correctly identified, which is not necessarily the case; and
2. The members of the >80 age group in reality have less social contacts than other age groups. Thus, they have less chance of exposure to the virus than younger population.

Lower bound estimation results are summarized in Table 1 for different regions. It is observed from the table that the ratio of lower bound estimate of positive case to the confirmed cases, for the two countries that have lower CFRs, namely South Korea and Germany, is very close to 1.0. This is while this ratio is much greater than 1.0, for the two countries that have higher CFRs, namely Italy and Spain. Further, Fig. 2 juxtaposes the age-distribution of the resulting lower bound estimate with that of the confirmed cases and region’s population for Italy, South Korea, Germany, and Spain. Interestingly, South Korea and Germany, which have lower CFRs, show more similarity between the age-distributions of estimated positive cases and population. On the other hand, in Italy and Spain with higher CFRs, the discrepancy between the two distributions is more strongly pronounced.

**Table 1:** Juxtaposition of the age-distribution of the resulting lower bound estimation with that of the confirmed cases and region’s population for 6 different countries

| Country | Collection Date | Confirmed Cases (CC) | Lower Bound Estimate (LBE) | LBE/CC | CFR%<sup>+</sup> | Median Age [26] |
| --- | --- | --- | --- | --- | --- | --- |
| Italy | March 31 | 94,088 | 237,086 | 2.52 | ~14.2 | 46.5 |
| Spain | April 13 | 119,994 | 360,988 | 3.01 | ~12.2 | 43.9 |
| Germany | April 15 | 126,937 | 145,171 | 1.14 | ~2.6 | 47.8 |
| South Korea | April 15 | 10,591 | 13,203 | 1.25 | ~2.1 | 43.2 |

<sup>+</sup> Calculated with regression method [4]

**Figure 1:**
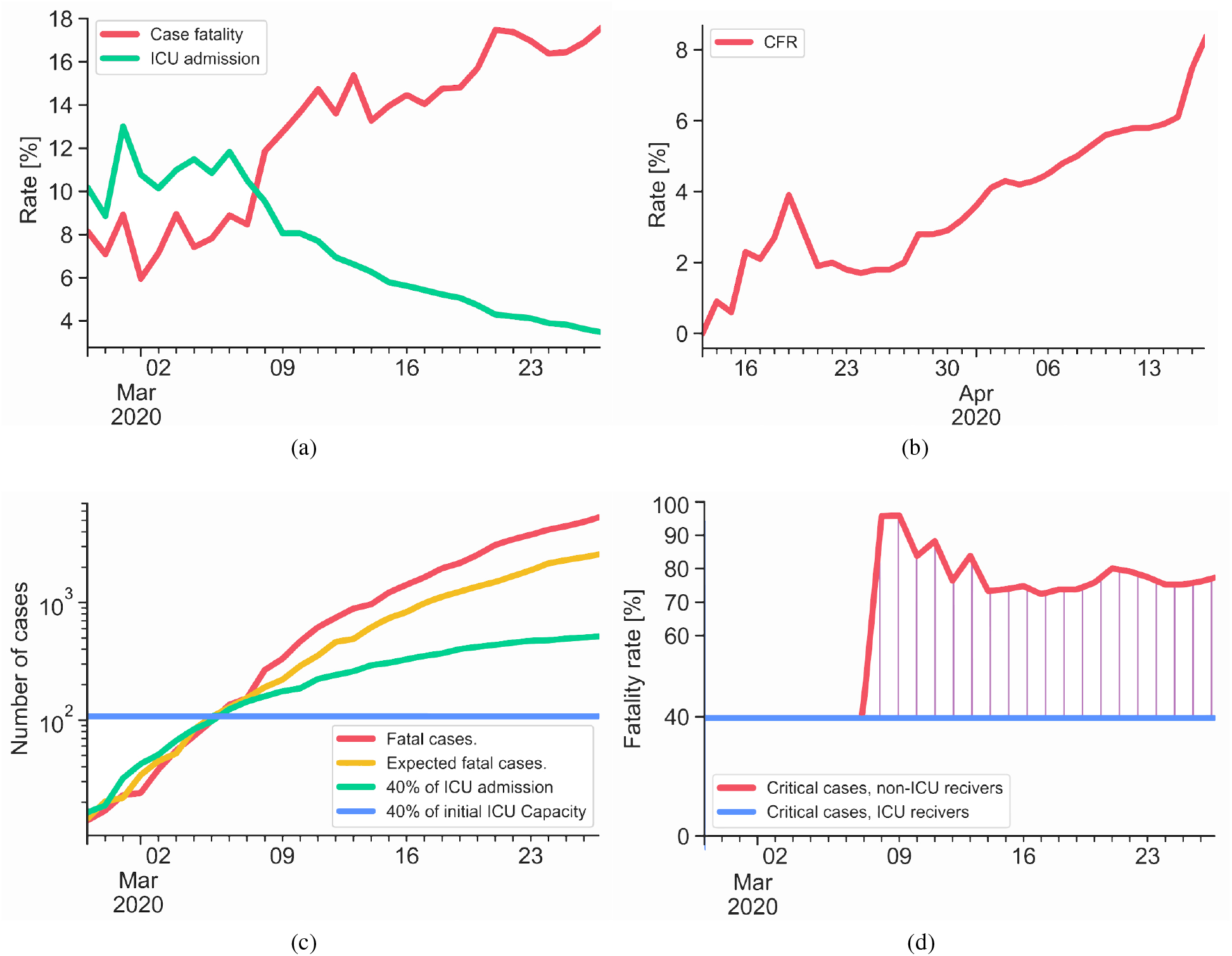
(a) Time-charts of CFR and percentage of critical case ICU-receivers for Lombardy, Italy; (b) Time-chart of CFR for New York; (c) Time-charts of CFR, expected CFR, 40% of intensive care receivers and 40% of initial health care capacity; (d) Comparison of fatality rate among ICU-receivers and non-ICU-receivers.

**Figure 2:**
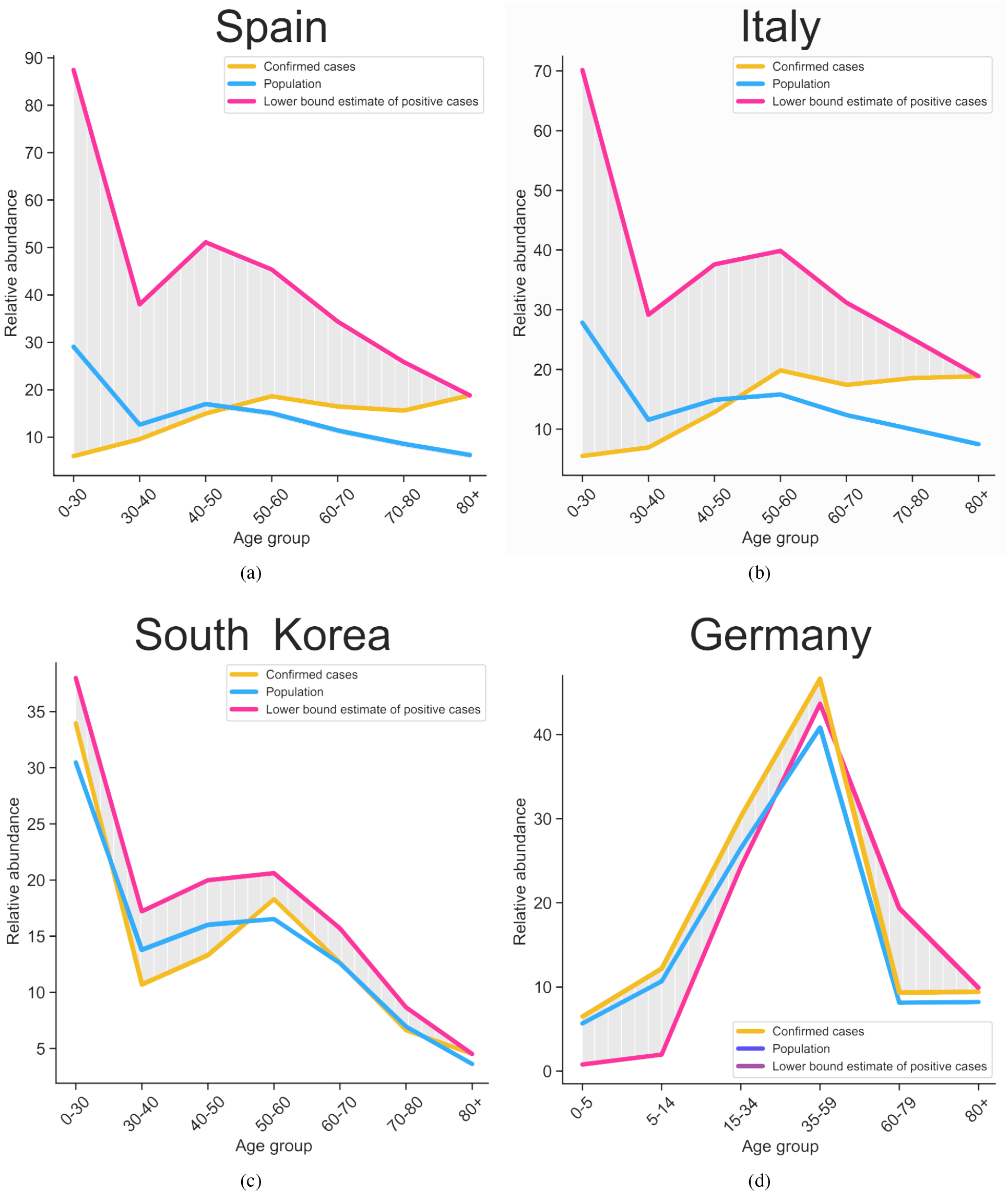
Juxtaposition of age-distribution of confirmed case, region’s population and estimated positive cases for (a) Spain (b) Italy (c) South Korea and (d) Germany.

### The effect of health care capacity on CFR

Realizing the fact that, access to ICU is necessary for patients with critical conditions [Ref: Booth and Stewart [22], Arabi, Murthy, and Webb [23]], we studied the effect of the number of available ICU beds on CFR. The analysis was performed on the data from two regions that were severely influenced by the pandemic, namely Lombardy and the New York State, as elaborated in the following.

### Lombardy

The region has the possession of about 18% of the total hospital beds in Italy. This is while, more than 35% of confirmed cases of Italy belong to Lombardy. Further, although Lombardy’s health care quality metrics are very similar to those of Italy’s average, the CFR for Lombardy is almost two times higher than the entire Italy excluding Lombardy (17.6% vs 9.7%, on March 27, calculated based on the number of confirmed cases).

To study how the health care capacity played a role in observing a higher CFR for Lombardy, Fig. 1a overlays the curves for CFR and intensive care rate, versus time. Here, the intensive care rate reflects the ratio of the number of patients, receiving intensive care, over the confirmed number of cases. On March 7, the intensive care rate shows a drop, which could be due to the fact that limit of health care capacity was reached, while the number of cases that needed intensive care continued to grow. Following that, one day after this drop, a sudden increase in CFR was observed for Lombardy. It is also observed that, starting on March 7, CFR surpassed intensive care rate, which means, from this date on, some patients that critically needed intensive care did not get a chance to receive it.

The effect of reaching maximum health care capacity on CFR could also be seen in Fig. 1c. Motivated by a report from the World Health Organization (WHO), which provided numerical percentages for mild, severe, critical and fatal cases, a best fit curve was obtained to overlay shifted curve for fatal cases and the curve for a fraction of critical cases. World Health Organization (WHO) provided numerical percentages for mild, severe, critical and fatal cases [24]. Motivated by this, we investigated the possibility of correlating the number of fatal cases to the number of cases that received intensive care. For this purpose, first, the fatal cases were shifted based on the phase lag value obtained from CFR calculations [4]. Next, a percentage factor that could correlate the shifted number of fatal cases to the number of intensive care cases was obtained. This was done by minimizing an error describing the difference between the two curves. The resulting percentage value was ∼40%. Accordingly, we compared, in the chart of Fig 1c, the numbers for fatality cases versus 40% of cases that received intensive care. In this chart, the horizontal line represents 40% of intensive care capacity. It is observed that, fatal cases and 40% of intensive care receivers, which fairly overlap until March 5, start to diverge on March 6, when the maximum capacity of intensive care is reached. The 40% factor used for intensive care capacity is for the normalization purpose, to assure that the numbers of intensive care receivers and intensive care capacity are compared on the same scale. Note that, the number of intensive care receivers continues to grow after it reaches the ICU cap, which implies that additional ICU beds have continuously been added but with a rate slower than required to accommodate all cases in need of it.

To quantify the importance of ICU beds in reducing fatality, the expected number of fatal cases in the hypothetical scenario that no cap existed on the number of ICU bed, was compared with the actual number of fatal cases. Such expected number of fatal cases was estimated using the number of confirmed cases and an average CFR value calculated from the days prior to reaching the cap of ICU beds. This CFR number was ∼8.45% as can be construed from Fig. 1a. The resulting expected number of fatal cases is shown in Fig. 1c, which follows the actual number of fatal cases, before ICU bed cap is reached, but remains below the actual number of cases ever since. The numbers for actual fatal cases, the expected fatal cases as well as the number of cases that receive intensive care are used to quantitatively estimate the effect of shortage of ICU beds on exacerbation of fatality rate. For this purpose, the fatality rate was estimated for critical cases who did not have access to standard ICU beds and was compared with that of those who had access to them, which are assumed to form ∼40% of the cases.

This result is presented in Fig. 1d. Note that, as opposed to the fatality rate for the group that had access to ICU beds, the fatality rate of the group that did not have access to ICU is graphed only after March 8. The reason for doing so is that, the latter group essentially was non-existing before March 8, as the cap of ICU beds was not reached yet. The fatality rate for the group, without access to ICU, spiked to ∼96% right after the cap was reached and slowly stabilized at ∼76%. This means that the chance of living for cases in need of intensive care, but not having access to it, was only about ∼24% as opposed to he ∼60% chance of living for the group that received the intensive care. The fatality rate of the group with no intensive care never reached 40%.

### New York

Based on an 82% occupancy rate for ICU beds in New York, as reported in Wunsch et al. [25], we have estimated the number of available ICU beds in New York to be 850. Similar to Lombardy’s case, by assuming a 40% fatality rate for critical cases who receive intensive care, it is estimated that the cap of health care is reached when the fatality cases are 340, which occurs on March 26. Figure 1b depicts the CFR for New York over time. Interestingly, it is evident, that on March 26, there is a sudden raise in CFR value, which supports the hypothesis that reaching health care cap immediately exacerbates CFR.

## Conclusion

In this work, we have proposed a method for estimating a lower bound for the number of positive cases by using the reported data on the oldest age group and the regions’ population distributions. The proposed estimation method improved the expected similarity between the age-distribution of positive cases and region’s population. Moreover, it was observed that regions with higher CFR show more discrepancy between the age-distribution of confirmed cases and region’s population. This discrepancy was quantified by calculating the error of confirmed cases against our estimated lower bound. Further, we have provided a quantitative measure for the impact of intensive care on the critical cases by comparing the CFR among those who did and did not receive intensive care. We showed that ∼40% of intensive care cases are fatal and that lack of access to ICUs immediately impacts the CFR. Our results showed that the chance of living among critical cases who don’t receive the intensive care remains less than half of ICU receivers (∼24% vs ∼60%). The proposed approach can serve as a generic tool for the community to conduct rigorous quantitative analysis on COVID-19 data from various regions. The resulting analyses provide insight for the policy makers regarding positive case distribution of the disease and the real impact of intensive care capacity on the fatality rate of COVID-19.

## Data Availability

All referred data is open access.

